# Disease-Modifying, Neuroprotective Effect of N-acetyl-L-leucine in Adult and Pediatric Patients with Niemann–Pick disease type C

**DOI:** 10.1101/2024.10.11.24315318

**Authors:** Marc Patterson, Uma Ramaswami, Aimee Donald, Tomas Foltan, Matthias Gautschi, Andreas Hahn, Simon Jones, Miriam Kolnikova, Laila Arash-Kaps, Julien Park, Stella Reichmannová, Mark Walterfarng, Pierre Wibawa, Marianne Rohrbach, Kyriakos Martakis, Tatiana Bremova-Ertl, P. Gissen

## Abstract

**Background:** The phase 3 randomized, placebo-controlled, crossover trial, IB1001-301, comparing N-acetyl-L-leucine (NALL) with placebo for the treatment of Niemann-Pick disease Type C (NPC) after 12 weeks met both its primary and secondary endpoints. In an open-label Extension Phase (EP) follow-up data have been obtained to evaluate the long-term effects of NALL for NPC. Here, we report on the safety and efficacy after 12 and 18 months of extended follow-up.

**Methods:** In the ongoing EP, pediatric and adult NPC patients received treatment with orally administered NALL 2-3 times per day in three tiers of weight-based dosing. The primary endpoint was the modified 5-domain Niemann-Pick disease type C Clinical Severity Scale (5-Domain NPC-CSS) (range 0-25 points; lower score representing better neurological status). Comparisons were made to the expected annual trajectory of decline (i.e. disease progression) on the 5-domain NPC-CSS established in published natural history studies. Analyses were also performed on exploratory endpoints including the 15-domain and 4-domain NPC-CSS and Scale for Assessment and Rating of Ataxia (SARA) scale.

**Results:** A total of 54 patients aged 5 to 67 years have been enrolled in the EP. After 12 months, the mean (SD) change from baseline on the 5-domain NPC-CSS was -0.32 (2.43) with NALL versus 1.5 (3.16) in the historical cohort (95% Confidence Interval, -3.11 to -0.53; p=0.007), corresponding to a 121% reduction in annual disease progression. After 18 months, the mean (SD) change was -0.067 (2.94) with NALL versus 2.25 (4.74) in the historical cohort (95% Confidence Interval, -4.17 to -0.46; p=0.017). The results of the 15-domain and 4-domain NPC-CSS were consistent with the primary analysis. The improvements in neurological signs and symptoms demonstrated in the Parent Study’s primary SARA endpoint were sustained over the long-term follow-up. NALL was well-tolerated, and no treatment-related serious AEs occurred.

**Conclusion:** In patients with NPC, treatment with NALL after 12 and 18 months was associated with a significant reduction in disease progression, demonstrating a disease-modifying, neuroprotective effect.

**Trial Registration Information:** The trial is registered with ClinicalTrials.gov (NCT05163288; registered 06-Dec-2021), EudraCT (2021-005356-10). The first patient was enrolled into the EP on 08-Mar-2023.

## Introduction

The IB1001-301 clinical trial was a randomized, double-blind, placebo-controlled clinical trial (hereafter referred to as the “Parent Study”) of N-acetyl-L-leucine (NALL) for the treatment of Niemann-Pick disease type C (NPC) ^1,2^. The trial enrolled pediatric and adult patients with NPC, a rare (incidence 1:100,000), progressive, debilitating, and prematurely fatal autosomal-recessive lysosomal storage disorder ^3^. In the trial, the mean (±SD) change from baseline on the primary Scale for the Assessment and Rating of Ataxia (SARA) endpoint total score was −1.97±2.43 points after 12 weeks of receiving NALL and −0.60±2.39 points after 12 weeks of receiving placebo (least-squares mean difference, −1.28 points; 95% confidence interval, −1.91 to −0.65; p<0.001), demonstrating an improvement in neurological signs and symptoms and functioning on NALL versus placebo. The trial also met all secondary endpoints. When patients received placebo, after having crossed over from NALL treatment, there was a deterioration in neurological status, further establishing that treatment with NALL affects neurological manifestations ^1^.

The IB1001-301 clinical trial served as the basis for the Marketing Approval of NALL (AQNEURSA**^™^**, levacetylleucine) by the US Food and Drug Administration (FDA) on the 24^th^ of September 2024. It is also the basis for an ongoing Marketing Authorization Application currently under review by the European Medicines Agency (EMA). Having demonstrated the benefits of treatment with NALL in the Parent Study, an extended open-label Extension Phase (EP) was planned to further investigate the long-term, neuroprotective effects of NALL treatment. Here, we report the results of this long-term follow-up after 12 and 18 months.

## Methods

### Standard Protocol Approvals, Registrations, and Patient Consents

Approval for the study (ClinicalTrials.gov identifier NCT05163288, EudraCT number 2021-005356-10) was obtained by National Regulatory Authorities in each country (US - Food and Drug Administration, UK - Medicines and Healthcare products Regulatory Authority, Germany - Federal Institute for Drugs and Medical Devices, Slovakia - Štátny ústav pre kontrolu liečiv, Switzerland - Swissmedic, Netherlands - Central Committee on Research Involving Human Subjects, Czech Republic - State Institute for Drug Control, and Australia - Therapeutic Goods Administration, and the applicable responsible central research ethics committees / institutional review boards for each center (Ethics Committee of Ludwig Maximilian University of Munich (21-1269;), National Institute of Child Diseases Bratislava Ethics Committee (EudraCT 2021-005356-10), Ethics Committee of General University Hospital in Prague (56/22 S-MEK), East Midlands – Derby Research Ethics Committee (1004498), Amsterdam UMC Locatie AMC Central Ethics Committee (NL79787.018.21), Mayo Clinic Institutional Review Board (22-001734), Emory University Institutional Review Board (STUDY00003227) Ethics Committee of the Canton of Bern, Switzerland (BASEC 2022-00638), and Victoria Human Research Ethics Committee (HREC/86167/MH-2022). Written informed consent was obtained for all study participants by the patient or, if applicable, their parent or legal representative. The trial is registered with ClinicalTrials.gov (NCT05163288; registered 06-Dec-2021), EudraCT (2021-005356-10). The first patient was enrolled into the EP on 08-Mar-2023.

### Participants

The IB1001-301 Parent Study was a randomized, placebo-controlled, crossover trial comparing orally administered NALL and placebo in patients aged ≥4 years with a diagnosis of NPC. To be eligible for the study, patients must have presented with clinical symptoms and signs referable to NPC, provided informed consent (or responsible person), and undergone a washout of any prohibited medications (N-acetyl-DL-leucine, N-acetyl-L-leucine, Sulfasalazine, Rosuvastatin) for 42 days before screening. The trial was approved by all respective ethics committees/ institutional review boards ^2^.

Patients who completed the final scheduled visit of the Parent Study (Visit 6) were eligible to continue into an open-label Extension Phase (EP) under the same trial protocol. The EP was conducted at the same trial sites, and with the same Investigators, as the Parent Study. Eligible participants were those who (a) completed the Parent Study Visit 6, (b) for whom the Investigator determined continued treatment with NALL may be in their best interest, and (c) who (or their legal representative) provided written informed consent to continue in the EP.

### Extension Phase Study Design

The EP is an open-label study. The trial consisted of a baseline visit, conducted in tandem with the last visit of the Parent Study (Visit 6). The EP visit was called “Visit 7” (4 patients had independent Visit 7 visits conducted 28, 42, 57, and 64 days after Visit 6 to accommodate each family’s scheduling requests). Following this baseline visit, patients received open-label treatment with NALL for a minimum of 1 year (365 +/-14 days). Visits occurred at 6 months (Visit 8, 180 +/- 14 days) and after 1 year (Visit 9, 365 +/- 14 days) (Figure 1). The EP is ongoing. A prespecified analysis was planned after all patients who participated in the Parent Study and continued into the EP had completed Visit 9.

**Fig 1.**
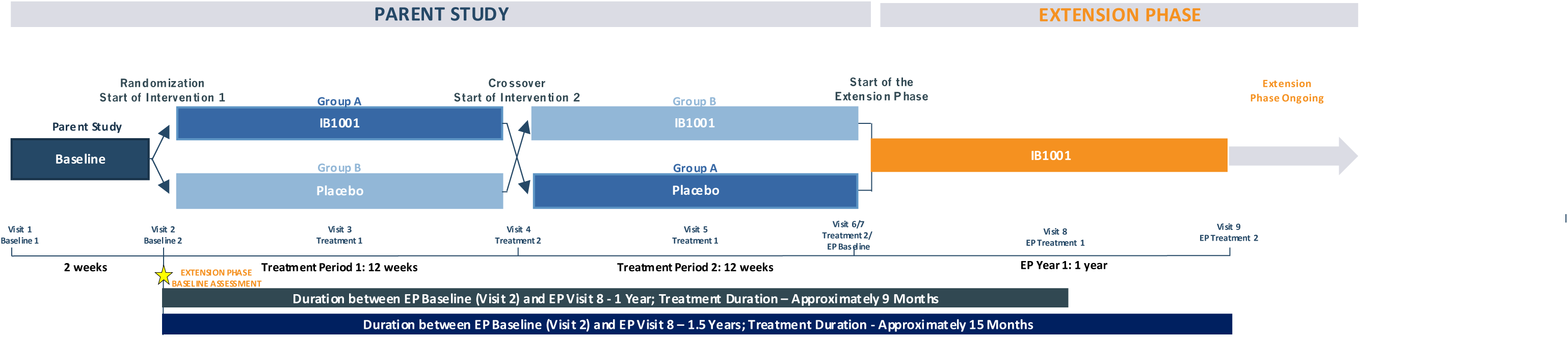
IB1001-301 Extension Phase Scheme. EP: Extension Phase

Given the placebo-controlled crossover design of the Parent Study, n=28 (52%) of patients were receiving NALL at EP Baseline Visit 7. Accordingly, it was pre-specified that the Parent Study Randomization Baseline Visit (Visit 2) was utilized as the “baseline” visit for the EP analysis. The approximate duration between Baseline (Visit 2) and Visit 8 was 12 months and 18 months between Baseline (Visit 2) and Visit 9. Over these durations, patients received treatment with NALL for approximately 9 months and 15 months, respectively (Figure 1).

Patients aged ≥13 years or aged 4-12 years weighing ≥35 kg received 4 g/day of orally administered NALL (granules in a sachet for suspension in 40 mL water, orange juice, or almond milk) three times per day (2 g in the morning, 1 g in the afternoon, and 1 g in the evening). Patients aged 4-12 years weighing <35 kg received weight-tiered doses two or three times per day based on an approximate total dose of 0.1 g/kg/day.

### Outcomes

The primary endpoint of the Extension Phase was the modified 5-domain Niemann-Pick disease type C Clinical Severity Scale (5-domain NPC-CSS), a five-item (Ambulation, Cognition, Fine motor skills, Speech, and Swallow) clinical rating scale ranging from 0–25, where 0 is the best neurological status and 25 the worst ^4^. Each domain is rated on a scale of 0–5. The 5-domain NPC-CSS is an abbreviated assessment tool originating from the 17-domain NPC-CSS developed specifically by Yanjanin et al. (2010) as a clinical outcome assessment to characterize and quantify disease progression in patients with NPC. The 17-domain NPC-CSS consists of 9 major domains (Ambulation, Cognition, Eye movement, Fine motor skills, Memory, Seizures, Speech, Swallow, Hearing) and of 7 modifiers (Behavior, Gelastic cataplexy, Hyperreflexia, Incontinence, Narcolepsy, Psychiatric and Respiratory, Auditory Brainstem). In this study, the widely used 15-domain NPC-CSS was utilized (which excludes the hearing and auditory brainstem response domains) ^3^. The 15-domain NPC-CSS has a total score for overall neurological status ranging from 0 (best) to 54 (worst) and was utilized as an exploratory endpoint.

The definitions of response with respect to 5- and 15-domain NPC-CSS scores were selected to measure deviations from the expected trajectory of disease progression established in the published natural history studies in patients with NPC. Yanjanin ^5^ first reported, based on a cross-sectional evaluation of 37 NPC patients, disease progression could be modelled on the 17-domain NPC-CSS by the following equation: SC_t0+x_ = SC_t0_ + 1.87x; where SC_t0_ is the initial score and SC_t0+x_ is the predicted future score after x years. Mengel ^6^ subsequently reported in a prospective observational study of 36 NPC patients, a mean (±SD) increase of 1.4± 2.9 on the 5-domain NPC-CSS (corresponding to an annualized progression rate of 1.5 points) and 2.7 ±4.0 on the NPC-CSS (excluding hearing) after 12 months.

In the above studies, a linear annualized increase (independent of age of disease onset, and similar in all patients) has been documented on the 5-domain / NPC-CSS scales, reflecting the progressive neurodegenerative nature of NPC. Therefore, a higher score indicates a clinical worsening (disease progression), while a lower 5-domain / NPC-CSS score indicates a clinical improvement and disease modification. A 0-point change represents a stabilization of disease progression, which is also a significant clinical benefit for rapidly progressive, neurodegenerative diseases such as NPC.

In a recent 12-month, double-blind, randomized placebo-controlled trial with the agent Arimoclomol, the mean progression after 12 months in the 16 patients receiving placebo was 2.15 points on the 5-domain NPC-CSS and 2.7 points on the NPC-CSS ^7^ (Table 2, 3). However, for the sake of this analysis, the conservative, validated linear natural history cohort values have been utilized as the basis for comparison, potentially underestimating the clinical deterioration in untreated patients.

Exploratory endpoints included the Scale for the Assessment and Rating of Ataxia (SARA), an eight-item (gait, stance, sitting, and speech disturbance, as well as the finger-chase test, the nose-to-finger test, the fast-alternating-hand-movements test, and the heel-along-shin slide test) clinical rating scale that incorporates functional assessments of gait, balance, speech, fine motor function, and upper/lower extremity function; scores range from 0 to 40, with lower scores indicating better neurological status ^8^. The 4-domain NPC-CSS (the 5-domain NPC-CSS with rescored swallow domain and excluding the cognition domain), which served as the basis for the FDA marketing authorization of the combination therapy Arimoclomol and Miglustat was also analyzed ^9^.

Safety assessments included monitoring for adverse events (whereby the site investigators or their delegates assessed the relation of the event to NALL), clinical laboratory testing and full pharmacokinetic sampling, physical examination, evaluation of vital signs, and electrocardiography.

### Statistical analysis

The number of patients entering the Extension Phase was determined by the number of patients completing the parent study (Visit 6) and who consented to participate in the open-label follow-up with NALL. The primary endpoint was defined as the numerical difference of the 5-domain NPC-CSS value for patients treated with NALL from baseline (Visit 2) versus 12 months (Visit 8) and 18 months (Visit 9) evaluated against benchmark annual mean rates of progression from the historical cohorts under the standard of care of 1.5 points annually for the 5-domain and 1.87 points annually for the 15-domain NPC-CSS.

An independent-sample t-test at a two-sided 5% significance level was used to test the null hypotheses that the mean change from the Extension Baseline on the 5-domain NPC-CSS and the 15-domain NPS-CSS score is equal to or greater than the 12 months or 18 months change expected in the natural history cohorts. The mean and standard deviation for the 18-month historical control cohort was modelled based on the formulas for the annualized increase. Point estimates and 95% confidence intervals of the mean difference are presented. The mean change from the Extension Baseline and 95% confidence intervals are computed for the exploratory endpoints: 4-domain NPC-CSS and SARA. A one-sample t-test at two-sided 5% significance level was used to test whether the change in SARA from Extension Baseline differed from zero after 12 and 18 months no historical data exists on the 4-domain NPC-CSS, this endpoint was reported descriptively.

For the 4-, 5-, 15-domain NPC-CSS and the SARA score, descriptive tables are presented with data available from all published or publicly presented previous natural history cohorts and clinical trial cohorts. The data is presented for each scale at 12 months (consistent with what is available in the literature and public domain) (see Tables 2-5). The mean and SD (if available) for each cohort are presented, along with the mean difference from the IB1001-301 NALL EP cohort. If available, for clinical trial cohorts treated with drug therapies, the data with and without miglustat on each scale at 12 months is presented.

The primary analysis was performed according to the modified intention-to-treat (mITT) principle, used to estimate the treatment effect regardless of discontinuation and to provide a perspective of the treatment effect across the entire population. The Extension Phase Modified Intention to Treat analysis set (mITTe) consisted of all patients aged 4 years and older who receive at least one dose of study drug (N-Acetyl-L-Leucine) in the Extension Phase, and with NPC-CSS scores at Extension Analysis Baseline (Visit 2)) and during the Extension Phase Treatment Period I (Visit 8 or Visit 9).

The Safety Analysis Extension Phase Set (SAFe) consisted of all patients who received at least one dose of study drug in the Extension Phase. The safety, integrity, and feasibility of the trial were monitored by an independent data safety monitoring board (DSMB) consisting of three independent, non-participating members (including two clinicians and a statistician).

### Data Availability

All authors had full access to all the data in the study and had final responsibility for the decision to submit for publication. The study Sponsor, IntraBio Inc. is committed to providing qualified scientific researchers appropriate access to anonymized data and clinical study information from the company’s clinical trials for the purpose of conducting legitimate scientific research. Requests for specific data will be considered along with the rationale, description of use need, and clinical value of the proposed analysis. IntraBio supports an approach to sharing data that responsibly reflects the interests of all parties involved in clinical trials, including protecting the rights and privacy of trial participants, the innovator’s intellectual property rights, and other incentives for innovation, and as such, will evaluate requests for sharing company clinical trial data with qualified external scientific researchers. Requests to access the data from this clinical trial may be made at. Data will be made available for request after product approval in the United States and European Union, after product development is discontinued, or as otherwise required by law or regulation. There are circumstances that may prevent the Sponsor from sharing the requested data as the product is investigational at this time.

## Results

### Study Population

Between 08 March 2023 and 15 August 2023, 54 patients were enrolled in the Extension Phase (aged 5 to 67 years). The demographic and baseline clinical characteristics of the enrolled patients are presented in Table 1. Fifty patients qualified for the primary Modified Intention-to-Treat Extension Phase (mITTe) analysis set (92.6%), which included all patients dosed who had an NPC-CSS score at baseline and at least one Extension Phase treatment visit (Visit 8 or Visit 9). Three patients an NPC-CSS score at baseline due to accidentally missed assessment, and thus were not included; one patient was withdrawn after Visit 7 due to withdrawal of consent.

**Table 1.**
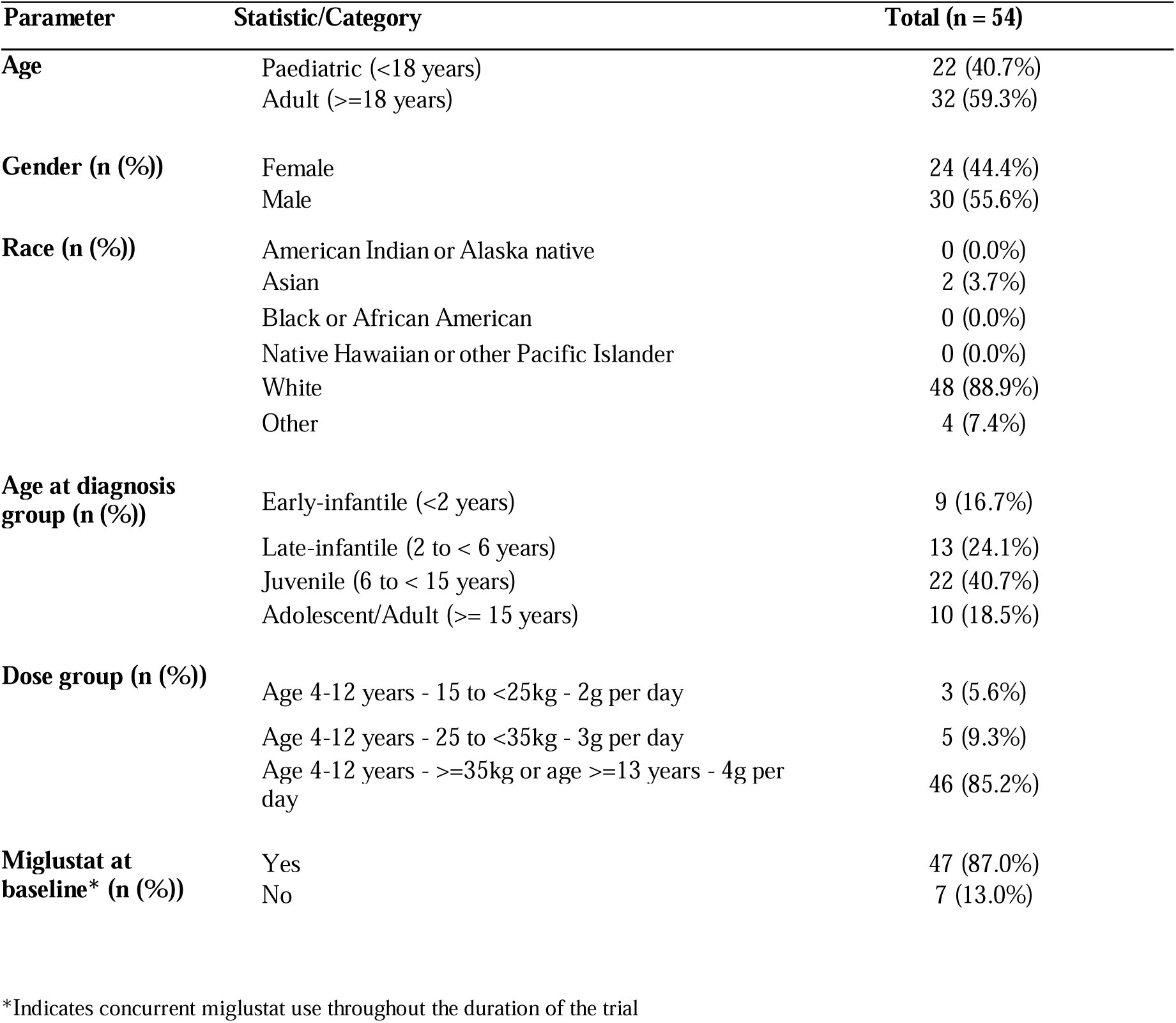
IB1001-301 Extension Phase (EP) Demographics and Baseline Characteristics.

| **Parameter** | **Statistic/Category** | **Total (n = 54)** |
| --- | --- | --- |
| **Age** | Paediatric (<18 years) | 22 (40.7%) |
|  | Adult (>=18 years) | 32 (59.3%) |
| **Gender (n (%))** | Female | 24 (44.4%) |
|  | Male | 30 (55.6%) |
| **Race (n (%))** | American Indian or Alaska native | 0 (0.0%) |
|  | Asian | 2 (3.7%) |
|  | Black or African American | 0 (0.0%) |
|  | Native Hawaiian or other Pacific Islander | 0 (0.0%) |
|  | White | 48 (88.9%) |
|  | Other | 4 (7.4%) |
| **Age at diagnosis group (n (%))** | Early-infantile (<2 years) | 9 (16.7%) |
|  | Late-infantile (2 to < 6 years) | 13 (24.1%) |
|  | Juvenile (6 to < 15 years) | 22 (40.7%) |
|  | Adolescent/Adult (>= 15 years) | 10 (18.5%) |
| **Dose group (n (%))** | Age 4-12 years - 15 to <25kg - 2g per day | 3 (5.6%) |
|  | Age 4-12 years - 25 to <35kg - 3g per day | 5 (9.3%) |
|  | Age 4-12 years - >=35kg or age >=13 years - 4g per day | 46 (85.2%) |
| **Miglustat at baseline\* (n (%))** | Yes | 47 (87.0%) |
|  | No | 7 (13.0%) |

\*Indicates concurrent miglustat use throughout the duration of the trial

**Table 2.**
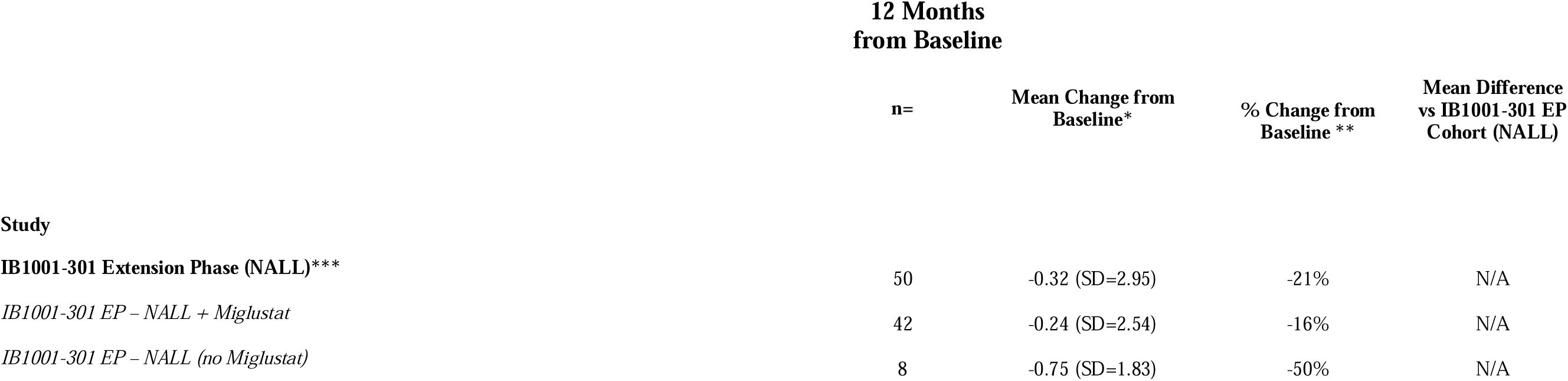
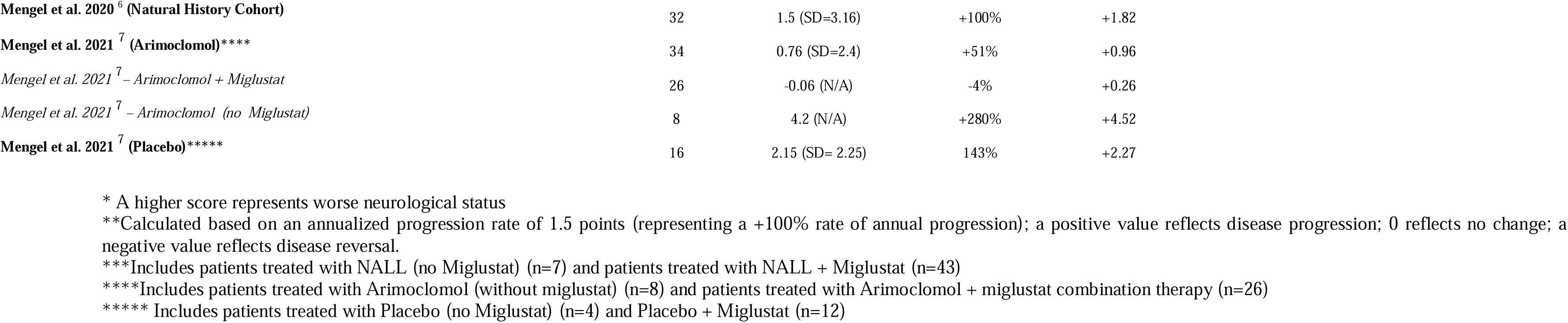
Comparison of the 5-Domain NPC-CSS while receiving NALL with Historical & Clinical Trial Cohorts with NALL.

|  | 12 Months<br>from Baseline |  |  |  |
| --- | --- | --- | --- | --- |
|  | n= | Mean Change from<br>Baseline* | % Change from<br>Baseline ** | Mean Difference<br>vs IB1001-301 EP<br>Cohort (NALL) |
| **Study** |  |  |  |  |
| **IB1001-301 Extension Phase (NALL)\*\*\*** | 50 | -0.32 (SD=2.95) | -21% | N/A |
| *IB1001-301 EP – NALL + Miglustat* | 42 | -0.24 (SD=2.54) | -16% | N/A |
| *IB1001-301 EP – NALL (no Miglustat)* | 8 | -0.75 (SD=1.83) | -50% | N/A |
| **Mengel et al. 2020<sup>6</sup> (Natural History Cohort)** | 32 | 1.5 (SD=3.16) | +100% | +1.82 |
| **Mengel et al. 2021<sup>7</sup> (Arimoclomol)\*\*\*\*** | 34 | 0.76 (SD=2.4) | +51% | +0.96 |
| *Mengel et al. 2021<sup>7</sup> – Arimoclomol + Miglustat* | 26 | -0.06 (N/A) | -4% | +0.26 |
| *Mengel et al. 2021<sup>7</sup> – Arimoclomol (no Miglustat)* | 8 | 4.2 (N/A) | +280% | +4.52 |
| **Mengel et al. 2021<sup>7</sup> (Placebo)\*\*\*\*\*** | 16 | 2.15 (SD= 2.25) | 143% | +2.27 |

\* A higher score represents worse neurological status
\*\*Calculated based on an annualized progression rate of 1.5 points (representing a +100% rate of annual progression); a positive value reflects disease progression; 0 reflects no change; a negative value reflects disease reversal.
\*\*\*Includes patients treated with NALL (no Miglustat) (n=7) and patients treated with NALL + Miglustat (n=43)
\*\*\*\*Includes patients treated with Arimoclomol (without miglustat) (n=8) and patients treated with Arimoclomol + miglustat combination therapy (n=26)
\*\*\*\*\* Includes patients treated with Placebo (no Miglustat) (n=4) and Placebo + Miglustat (n=12)

Visit 8 occurred approximately 1 year after the Baseline Visit (Visit 2) during which patients received treatment with NALL for approximately 9 months (mean duration of 268 [min 233, max 287] days out of mean 354 days [min 317, max 371]). Visit 9 occurred approximately 18 months after the Baseline Visit (Visit 2) during which patients received treatment with NALL for approximately 15 months (mean duration of 453 [min 435, max 556] days out of 538 days [min 520, max 633]) (Figure 1). The EP remains ongoing for additional years of long-term follow up.

### Efficacy

#### Primary Endpoint

The mean (± SD) baseline (Visit 2) on the 5-domain NPC-CSS was 11.04 (±4.70). After 12 months (Visit 8), the mean change from baseline was -0.32 ± 2.43 points with NALL versus 1.5 ± 3.1 points in the historical cohort [mean difference 1.82 points; 95% confidence interval [CI], -3.11 to -0.53; p=0.007]. After 18 months (Visit 9), the mean change from baseline was -0.067 ± 2.94 with NALL versus 2.25 (4.74) in the historical cohort [mean difference 2.32 points; 95% confidence interval [CI], -4.17 to -0.46; p=0.017). Table 2 / Figure 2 reports and depicts the change in the 5-domain NPC-CSS with NALL compared with all published historical cohorts as well as clinical trial cohorts.

**Fig. 2:**
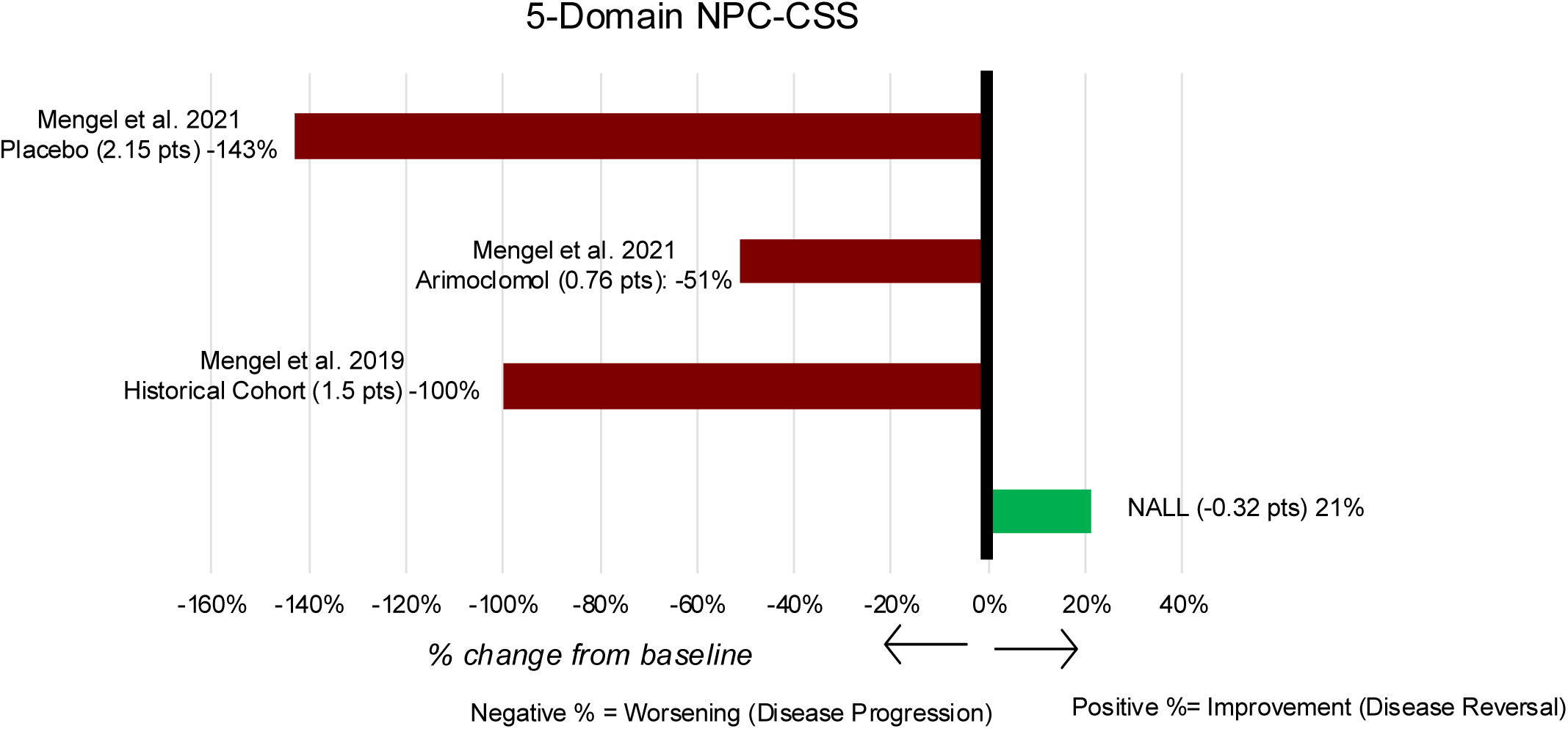
Bar plot for 5-Domain NPC-CSS Total Score versus Published Historical Cohorts & Clinical Trial Cohorts. Percentage calculated based on the annualized progression rate of 1.5 points (representing a -100% rate of annual progression); a negative value reflects disease worsening/progression; 0 reflects no change (disease stabilization); and a positive value reflects disease improvement.

#### Exploratory Endpoints

The mean (± SD) baseline (Visit 2) score on the 15-domain NPC-CSS was 18.22 (±7.14). After 12 months (Visit 8), the mean change from baseline was -0.06 ± 3.27 points with NALL versus 1.87 ± 1.09 points in the historical cohort [mean difference -1.93; 95% confidence interval [CI], -2.90 to -0.96; p<0.001]. After 18 months (Visit 9), the mean change from baseline was 0.29 ± 4.67 with NALL versus 2.81 ± 1.64 in the historical cohort [mean difference -2.52; 95% confidence interval [CI], -3.98 to -1.05; p=0.001]. Table 3 reports the change on the 15-domain NPC-CSS with NALL compared with all published historical cohorts as well as published clinical trial cohorts.

**Table 3.**
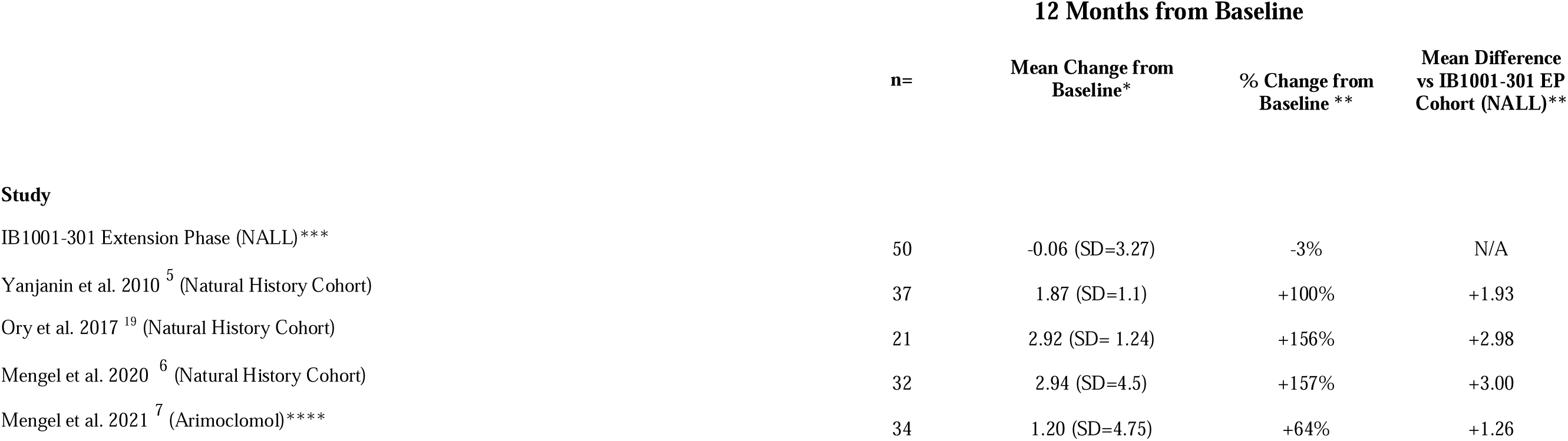
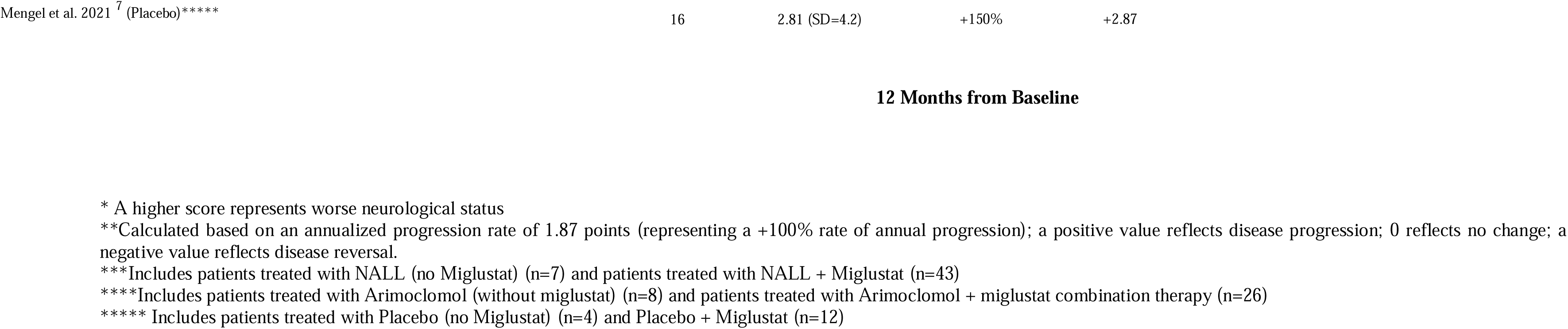
Comparison of the NPC-CSS while receiving NALL with Historical Cohorts & Clinical Trial Cohorts.

The mean (± SD) baseline (Visit 2) score on the 4-domain NPC-CSS was 8.10 (±3.56). After 12 months (Visit 8), the mean change from baseline was -0.62 ± 1.78 points with NALL. After 18 months (Visit 9), the mean change from baseline was -0.33 ± 2.28 NALL. Table 4 reports the change on the 4-domain NPC-CSS with NALL compared with all publicly available cohorts.

**Table 4.**
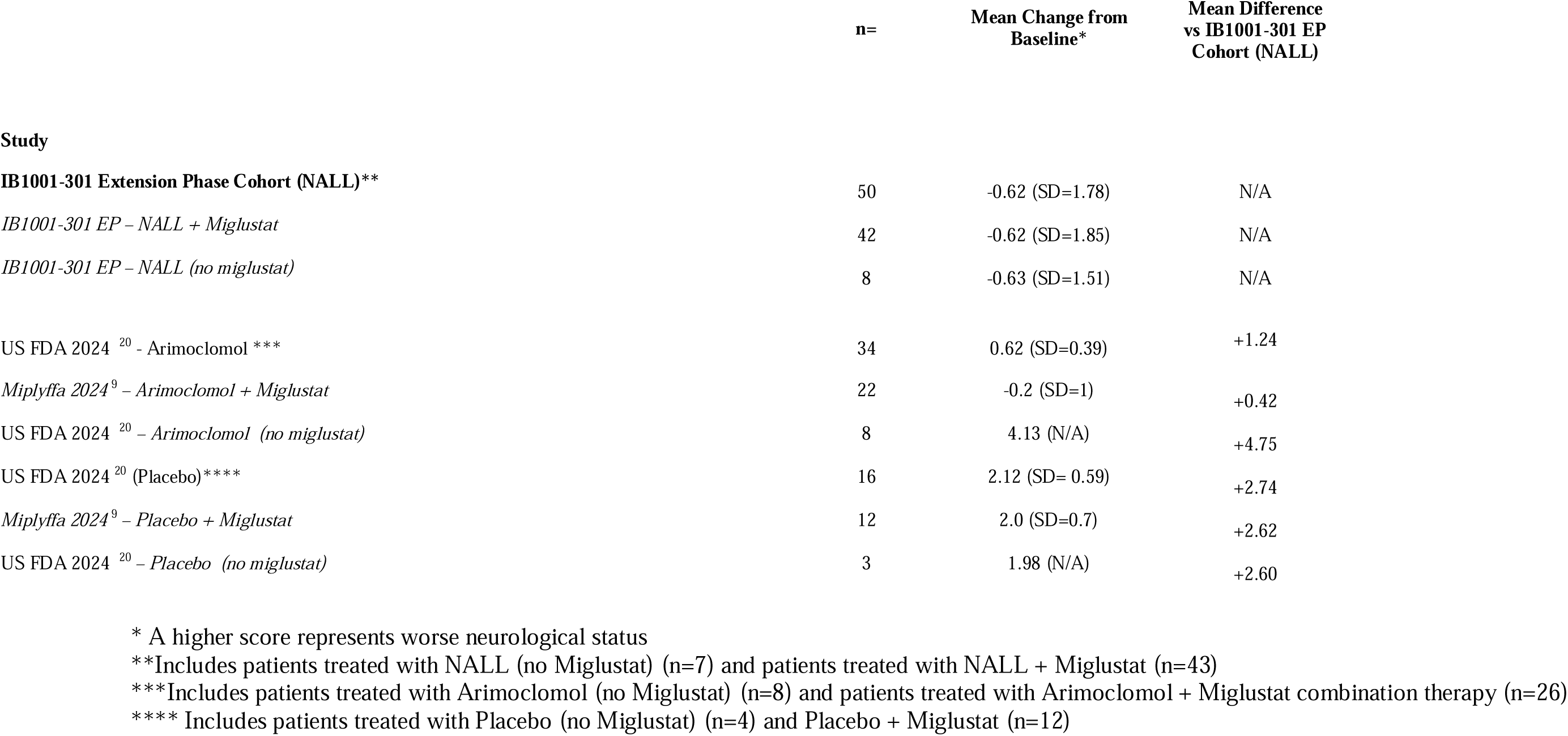
Comparison of the 4-Domain NPC-CSS while receiving NALL with Historical & Clinical Trial Cohorts with NALL.

**Table 5.**
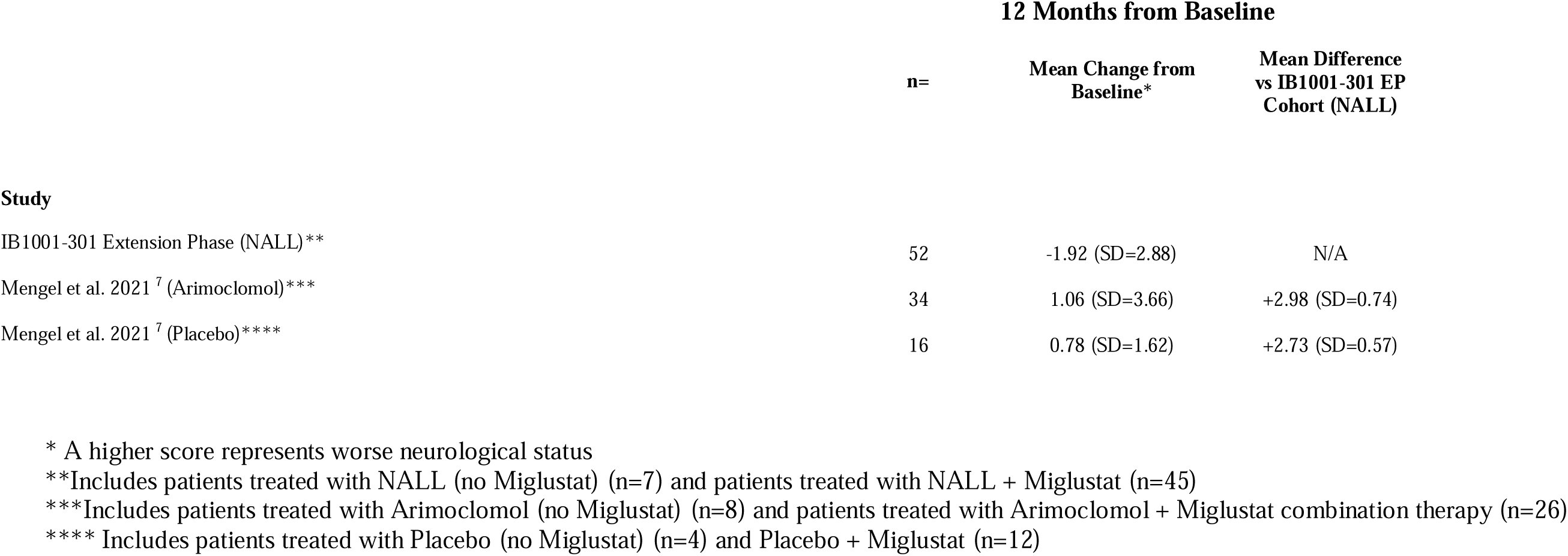
Comparison of the SARA score while receiving NALL with Clinical Trial Cohorts.

The mean (± SD) baseline (Visit 2) score on the SARA score was 15.80 (7.62). After 12 months (Visit 8), the mean change from baseline was -1.92 ± 2.88 points with NALL (95% confidence interval [CI], 1.12 to 2.72; p <0.001). After 18 months (Visit 9), the mean change from baseline was -1.67 ± 3.21 with NALL (95% confidence interval [CI], -2.60 to - 0.73, p(mean change = 0) <0.001). There was no significant difference between these values and those from the completion of the Parent Study; rather, the improvements on the SARA were sustained across long-term treatment. Table 5 reports the change on the SARA with NALL compared with all published cohorts.

#### Safety

The Treatment-Emergent Adverse Events (TEAEs) are shown in Supplementary Table 5. No TEAEs led to premature discontinuation of the trial. No TEAEs occurred in more than 10% of patients on NALL and no patients had TEAEs that were assessed as by the investigator as related to NALL.

No trial drug-related serious adverse events (SAEs) or deaths occurred. Results of plasma and urine tests, vital signs, and ECG recordings were normal or rated as clinically non-significant. Adherence to trial drug was high as shown by treatment compliance and the regular urine analyses for prohibited medication.

## Discussion

Here we present findings from the largest cohort of NPC patients treated in the frame of a long-term clinical trial. The major findings of this study are as follows: in adult and pediatric patients with NPC, treatment with NALL was associated with a 121% reduction in annual disease progression after one year compared to a natural history control cohort, on the 5-domain NPC-CSS primary endpoint, reflecting a significant improvement in the patient’s condition from 1-year prior. After 1 year, there was a reduction of -0.32 points from baseline (compared to a 1.5-point increase on the historical cohort) on a 25-point scale used to assess neurological status in multiple domains and disease progression in NPC, and a change of -0.067 after 1.5 years (versus the expected +2.25 point change). This improvement is to date the most significant of any agent formally investigated in NPC (Table 2, 3,4; Figure2).

In the randomized, double-blind Parent Study, NALL demonstrated a significant improvement in neurological signs and symptoms after 12 weeks. The improvement in neurological manifestations and status demonstrated with NALL versus placebo on the SARA scale were maintained after 12 and 18 months of treatment, demonstrating a statistically significant and clinically meaningful improvement ^10^ and the long-term effects of NALL. No new safety signals and no drug-related serious adverse events were observed during the follow-up, reinforcing the benign safety profile of the molecule.

The improvement on the 5-domain NPC-CSS scale with NALL is clinically meaningful according to the validation of the 5-domain NPC-CSS, which demonstrated that a 1-point worsening on the 5-domain NPC-CSS constitutes a clinically meaningful change for caregivers/parents and physicians (e.g., a 1[point change or greater represents a clinically meaningful transition reflecting loss of complex function and increased disability), and therefore, preventing a 2-point worsening would be a viable treatment goal [Patterson et al. 2021]. Treatment with NALL not only prevented worsening (e.g. halted disease progression) on the 5-domain NPC-CSS, but led to an improvement, demonstrating a neuroprotective and disease-modifying benefit. This disease-modifying benefit was present in patients who were not receiving Miglustat (considered the standard of care but not authorized as an NPC-indicated treatment in the United States) ^11^ (Table 2).

The findings from this Extension Trial are consistent with NALL’s Mechanism of Action (MOA). The acetylation of leucine makes it a substrate for ubiquitously expressed monocarboxylate transporters (MCTs), delivering supraphysiological levels of NALL relative to leucine into the cell’s cytoplasm ^12^. There, NALL is deacetylated yielding L-leucine and enters enzyme-controlled pathways to correct metabolic dysfunction. For example, in the NPC mouse model (*Npc1^-/-^),* NALL leads to changes in Krebs cycle flux, shifting glucose metabolism from lactate/lactate dehydrogenase (LDH) towards pyruvate/pyruvate dehydrogenase (PDH) dependency, resulting in a significant improvement in adenosine triphosphate (ATP) production ^13^. Knock-on effects of restoring more efficient ATP synthesis include mitigating dysfunctional lysosome fusion and trafficking, resulting in reduced accumulation of cholesterol, sphingosines, and glycosphingolipids ^13^. The culmination of restoring mitochondria and lysosomal function includes normalization of neuronal membrane potential, restoring cellular signaling ^14^, and dampening of neuroinflammation leading to an overall reduction in neurodegeneration ^13,15–17^.

The disease-modifying effects of NALL have been clearly demonstrated in the NPC (*Npc1^-/-)^* mice, where pre-symptomatic treatment of *Npc1^-/-^* mice from 3 weeks of age delayed the onset of functional decline (gait abnormalities, motor dysfunction), the decline in general health, coat, and weight, slowed disease progression, and prolonged survival. These disease-modifying and neuroprotective effects were observed solely in animals exposed to the L-enantiomer (absent in animals treated with N-acetyl-D-leucine) ^13^.

Finally, the neuroprotective, disease-modifying findings are consistent with other non-clinical studies with the agent for common neurological conditions. For example, NALL restored autophagic flux and its neuroprotective function in the cortices of mice with Traumatic Brain Injury, leading to the attenuation of neuroinflammation and restriction of neuronal cell death, reflecting a neuroprotective effect ^16,17^. More recently, published observational studies in patients with prodromal Parkinson’s disease (REM sleep behavior disorder) utilized molecular imaging techniques in the brains of patients to show that acetyl-leucine stopped the decline of neuronal function at the cellular and molecular level, and reversed changes in the biomarker for prodromal alpha-synucleinopathies, preventing the clinical conversion to manifest Parkinson’s disease ^18^.

Limitations of the trial include the open-label design. Analyses were not adjusted for multiple comparisons. The data of the historical cohort were not made publicly available, thus it was not possible to match the historical cohort with the IB1001-301 EP patient population. Nevertheless, the IB1001-301 EP study population was significantly larger (31 vs. 54 patients with continued follow-up). However, due to the inclusion criteria of the Parent Study, the EP did not include patients aged <4 years, asymptomatic patients, or patients with advanced disease who would not be able, or reliably able, to complete functional assessments.

In conclusion, this study provides additional evidence of the benefits of NALL treatment. NALL represents a new class of therapeutic agent that acts as a metabolic modulator, rebalancing dysregulated energy metabolism and is pleiotropic in its effects. NALL is the first and only compound that has demonstrated both rapid improvements in neurological manifestations, as well as a long-term, disease-modifying, neuroprotective effect for patients with NPC. In light of its efficacy and safety profile, NALL should be considered a foundational and cornerstone therapy for the treatment of NPC. In addition, given the broad applicability of its mechanism, NALL should continue to be investigated for the treatment of a plethora of rare and common neurological disorders.

